# Sibling Experiences of Adverse Childhood Experiences: A Scoping Review

**DOI:** 10.1101/2022.02.04.22270452

**Authors:** Ben Donagh, Julie Taylor, Muna al Mushaikni, Caroline Bradbury-Jones

## Abstract

Adverse childhood experiences (ACEs) are traumatic events during childhood known to affect health and wellbeing across the life-span. The purpose of this scoping review was to understand what we currently know about the experiences of siblings living with ACEs. Sibling relationships are unique, and for some the most enduring we experience. These relationships can be categorised by love and warmth, however, can also be a point of escalating conflict and problems. This scoping review was conducted following Arksey and O’Malley’s (2005) methodological framework, complemented by the PAGER framework (Bradbury-Jones *et al*, 2021), offering a structured approach to the review’s analysis and reporting through presenting the **P**atterns, **A**dvances, **G**aps, and **E**vidence for practice and **R**esearch. In June 2020 we searched 12 databases, with 11,469 results. Articles were screened for eligibility by the review team leaving a total of 148 articles meeting the inclusion criteria. Findings highlighted five main patterns: (1) the influence of birth order (older siblings shielding younger); (2) the influence of sibling relationships (lack of research exploring sibling types outside of biological siblings); (3) identifying siblings experiencing ACEs (when one sibling experiences adversity, it is likely that their other siblings also do, or experience vicariously); (4) siblings who cause harm (siblings harming other siblings is often normalised and minimised, especially by parents); (5) focus on individual ACEs (the majority of studies explore ACEs in isolation). Our findings suggest future research would benefit from an increase in theoretical understanding and exploration of different types of sibling relationships (full, step, half).

## Background/ Context

Adverse childhood experiences (ACEs), of which there are most commonly 10, are traumatic events during childhood that are known to affect health and wellbeing across the life-span (Crouch et al., 2019; Felitti et al., 1998; Hargreaves et al., 2017; Leban & Gibson, 2019). There is not one fully agreed list of ACEs (Finkelhor, 2020). Depending on a study’s methodological approach, definitions and the number of ACEs measured can vary, with the first study conducted in the 1990s measuring only seven ACEs (Felitti et al., 1998; Manyema & Richter, 2019). The second wave of studies updated this list to a total of ten within three categories, which now appear to be the most commonly used:

- abuse (emotional abuse, physical abuse, sexual abuse),
- neglect (emotional neglect, physical neglect)
- and household dysfunction (domestic violence and abuse, substance abuse, mental illness, parental separation or divorce, incarceration) (Hargreaves et al., 2017; Manyema & Richter, 2019).

There does however continue to be variation between studies; a 2016 National Survey of Children’s Health in the United States chose to only measure nine ACEs, two of which were racial/ethnic mistreatment and economic hardship (Crouch et al., 2019). There are ongoing debates around which ACEs should be measured and the implications or limitations of studies when they do not include stressors that effect a child’s wellbeing; an example of this being poverty (Hughes & Tucker, 2018).

Variations amongst studies are also seen with their approach to measuring ACEs. Blum, Li, & Naranjo-Rivera, (2019) identified the four most commonly used approaches: (1) Cumulative ACEs scores, (2) Weighting individual ACEs, (3) Weighting ACEs by subgroup, and (4) ACEs typology.

1. Cumulative ACEs score is when the number of unique ACEs are added together and no consideration is given to frequency, duration or intensity (Blum, Li, & Naranjo-Rivera, 2019).
2. Weighting individual ACEs involves giving consideration to particular characteristics of each ACE and these being weighted accordingly; events that are more recent, severe or frequent are weighted higher (Blum, Li, & Naranjo-Rivera, 2019).
3. Weighting ACEs by subgroup consists of ACEs being grouped into categories and each category being weighted rather than the individual ACE, creating a hierarchy of severity (Blum, Li, & Naranjo-Rivera, 2019).
4. ACEs typology involves clustering ACEs together when assessing against them, such as ‘low ACEs’, ‘household dysfunction’, ‘emotional ACEs’, and ‘high/multiple ACEs’ (Blum, Li, & Naranjo-Rivera, 2019).

Felitti et al., (1998) used the ACEs typology approach, asking a total of eight questions within the category of childhood abuse and nine questions within the category of household dysfunction. Respondents were defined as exposed to a category if they responded “yes” to one or more of the questions in that category (Felitti et al., 1998).

The prevalence of ACEs has been discussed at length, with the original ACE study finding one in four adults to report three or more ACEs (Felitti et al., 1998). Later studies have found them to be somewhat more prevalent within particular populations such as children in care and children within the welfare system (Bramlett & Radel, 2014; Hargreaves et al., 2017). Correlations have also been identified with regards to the number of ACEs individuals are likely to encounter; Duke et al (2010) assert that individuals who report at least one ACE are likely to report experiencing others, with Baglivio & Epps (2016) strengthening this by finding 67% of their participants who were exposed to one ACE had also been exposed to four or more (Leban & Gibson, 2019).

Studies have found ACEs impact a child’s social and emotional development as well as causing poor health across their life course, such as having a greater risk of poor physical or mental health, chronic disease, and cancer (Blum, Li & Naranjo-Rivera, 2019; Choi, Wang, & Jackson, 2019; Crouch et al., 2019). Associations have also been found between ACEs and health harming behaviours such as drug use and smoking at an early age alongside developing depression and anxiety, and also premature death (Brown et al., 2009; Chapman et al., 2004; Crouch et al., 2019; Dube et al., 2003). Impact has also been documented around exposure disrupting healthy brain development in childhood (Crouch et al., 2019; Garner, 2013; Shonkoff et al., 2012; Shonkoff, Boyce, & McEwen, 2009).

The identification of ACE exposure can inform early interventions, thus potentially mitigating the long-term impact they can have, however limitations have been found with this process (Crouch et al., 2019). The lack of consistency in the number of ACEs measured and methodological approaches make it more difficult to create a consistent understanding between studies (Hargreaves et al., 2017; Manyema & Richter, 2019). In addition to this, most commonly the study of ACEs is undertaken retrospectively, relying on the recall of adults around exposure in childhood meaning potential information biases can be a major threat (Crouch et al., 2019; Anda et al., 2006; Felitti et al., 1998). Exploring ACEs contemporaneously with children and young people, rather than retrospectively as adults, has been found to improve the ability of services and caregivers to mitigate the exposure and impact, reducing the likelihood of poor outcomes (Crouch et al, 2019).

### Sibling relationships

Compared to other family relationships, sibling relationships are argued to be understudied despite being the longest-lasting relationship is most people’s lives (Gilligan, Stocker & Conger, 2020). As many as 85-90% of children are reported to grow up with at least one brother or sister (Milevsky, 2011; Tippett & Wolke, 2015). Siblings may or may not be blood related, and the definition of siblings can vary between studies. Kiselica and Morrill-Richards (2007 p.149) provide a useful outline of the different types of sibling relationships. These relationships include: “biological siblings, (share both parents), half-siblings (one parent in common), step-siblings (connected through marriage of parents), adoptive siblings, foster siblings (joined through a common guardian), or fictive siblings (united by emotional bond)”. Others (Bass et al, 2006) also recognise children who had been living together in the same family and had assumed the role of siblings for two or more years.

The sibling relationship is unique, and for some can be one of the most enduring relationship we have, starting at birth and continuing until death. Siblings can provide an important source of support and play a vital role in an individual’s wellbeing (Davies 2015; Edwards et al 2006 Exley, 2021; Yucel & Yuan, 2015). These relationships can be categorised by love and warmth, providing security and the opportunity to develop social abilities and self-identity (Davies 2015; Edwards et al 2006). However, sibling relationships can also be a point of escalating conflict and problems (Buist, Deković, & Prinzie, 2013). Some sibling relationships may be ingrained with rivalry and conflict, with distance being introduced when they leave the parental home.

To our knowledge, there have been no scoping reviews of the literature on sibling experiences of ACEs. Our scoping review helps to address this gap. We aim to provide valuable insight that could help statutory bodies, service providers and policy makers develop effective intervention and/or prevention approaches, as well as inform the direction of future research around this topic.

## Methods

This scoping review was conducted following Arksey and O’Malley’s (2005) methodological framework of a five-stage approach: identifying the research question; searching for relevant studies; selecting studies; charting data; and collating, summarising, and reporting the results. This approach has been complemented by the PAGER framework (Bradbury-Jones *et al*, 2021) which offers a structured approach to the analysis and reporting of scoping reviews through presenting the Patterns, Advances, Gaps, and Evidence for practice and Research within the included articles. Articles within this scoping review were identified following the Preferred Reporting Items for Systematic Reviews and MetaAnalyses extension for Scoping Reviews (Moher *et al*, 2009; Tricco et al., 2018). This comprehensive approach enabled an exploration across multiple disciplines to answer one key research question: What do we currently know about the experiences of siblings when living with Adverse Childhood Experiences (ACEs)? (Peters et al., 2015).

### Search Strategy

The following combination of terms were used to identify articles by searching the abstract and title: adolescen* OR child* OR teen* OR “young people” OR “young person” AND sibling* OR brother* OR sister* OR step-brother* OR step-sister* OR half-brother* OR half-sister* OR twin* AND “adverse childhood experience*” OR “childhood trauma” OR trauma* OR abuse* OR “physical abuse” OR assault* OR attack* OR “emotional abuse” OR “sexual abuse” OR rape* OR groom* OR “physical neglect” OR neglect* OR “emotional neglect” OR “mental illness” OR “mental health” OR “incarcerated relative*” OR prison* OR incarcerat* OR arrest* OR “domestic violence” OR “domestic abuse” OR “marital violence” OR “intimate partner violence” OR “gender-based violence” OR “substance abuse” OR “substance misuse” OR drug* OR alcohol* OR addict* OR divorc*.

Following multiple initial searches to finalise terms and databases, final searches were conducted in June 2020 producing 11,469 articles from 12 databases: Consumer Health Database, CINAHL, EMBASE, Healthcare Administration Database, Medline, PILOTS, PubMed, ProQuest Central, PsychInfo, Web of Science, Applied Social Sciences Index & Abstracts (ASSIA) and the International Bibliography of the Social Sciences. Inclusion criteria for eligible articles were:

1. Considered siblings in the context of an ACE.
2. Adversity took place between birth and 18.
3. Written in English.
4. Published 2005-2019
5. Peer-reviewed, empirical articles only

No limit was set for the age of participants so as to include both studies which collect data from children and young people and also those which collect from adults retrospectively.

Duplicates were removed, leaving 6,331 articles to be screened by title and abstract. 4,350 articles were excluded by title and a further 1,645 by abstract, leaving 336 articles for full text reviews. Initially, we planned to include articles from 1995-2019 to capture all articles published since the first ACEs study was conducted, however this strategy identified more results than we had the resources to review. The timescale was reduced by 10 years, meaning the final agreed timescale was 2005-2019 and a further 84 articles were excluded at this stage. Ten percent of the remaining 252 were reviewed by abstract and title by two reviewers (insert later and by full text by another reviewer (insert later). Any discrepancies were discussed and resolved. A total of 104 articles were excluded following full-text reviews, leaving 148 articles within this scoping review. See PRISMAScR Diagram in Figure 1.

**Figure 1.**
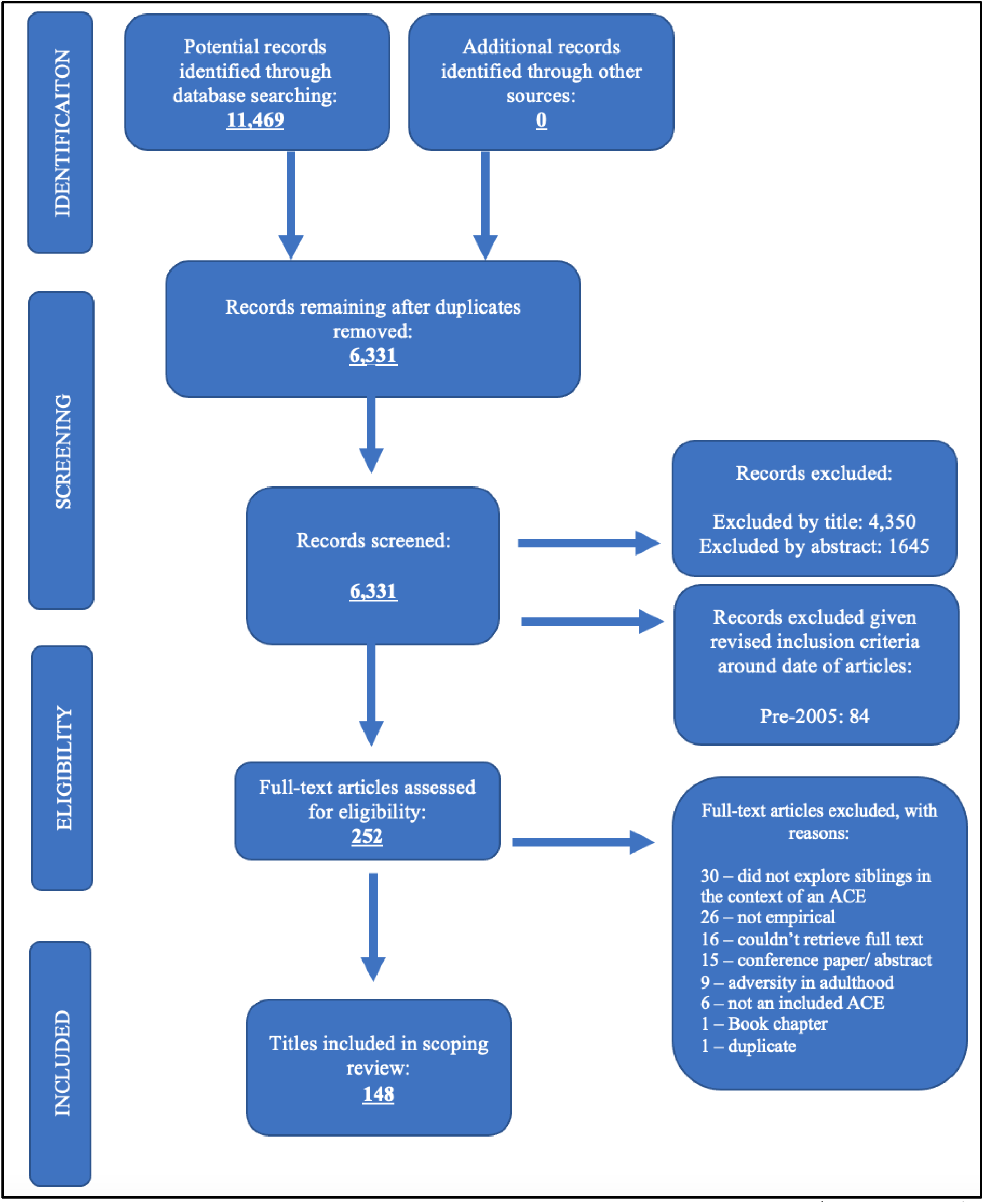
PRISMAScR Diagram.

A literature matrix was created, extracting the following data from each of the 148 included articles: author, year, title, country of origin, research method, theoretical input, sample size, sample demographic, type of ACE, whether the term ‘ACE’ was used, sibling type, whether siblings were the primary focus, and an overview of the article’s key findings. Analysis was completed using the PAGER Framework, starting with the identification of patterns (Bradbury-Jones et al, 2021). Patterns were identified in two stages; initially within each individual ACE category before being identified across multiple ACE categories. We created spider diagrams for each ACE as a visual tool to organise the data in a logical way. This process then enabled the analysis of data to identify Advances, Gaps, and Evidence for practice and Research.

## Findings

### Descriptive summary

Articles were located worldwide (25 countries) with the vast majority conducted in the USA (75 articles, 51%); a significant jump from the second highest being the United Kingdom (UK) at 17 articles (11%). Given the geographical reach, by default diversity will have been a consideration within some articles, based upon where they were conducted. However, the explicit discussion of diversity appears to be lacking. Within our review we have identified that age and gender commonly appear to have been considered within many articles, however other protected characteristics are not so commonly discussed.

The articles were clustered by childhood adversity type: physical abuse (27%; 40/148), emotional abuse (1%; 2/148), sexual abuse (20%; 30/148), neglect (1%; 2/148), mental illness (6%; 8/148), divorce (11%; 16/148), incarceration (4%; 6/148), substance abuse (7%; 10/148), domestic violence (9%; 14/148), ACE (8%, 12/148), maltreatment (6%; 8/148); see Figure 2. Disparities between the terminology and definitions used meant decisions were made regarding which ACE category some articles were grouped within. Articles which explored broad categories of *‘trauma’* or *‘child abuse’* (Foroughe & Muller, 2014; Heins *et al*, 2011; Lo, Lau & Yu, 2017) or explored multiple but not all of the ACEs included in this review (Hindle, 2007; Wolfe, 2016) were included in the ACE grouping. Also, the small number of articles exploring neglect (n=2) did not differentiate between physical and emotional neglect, so these have been grouped together within the overarching category of *‘neglect’*. A category of *‘maltreatment’* was introduced to the review following a number of articles (n=8) which used this term to cover a combination of abuse and neglect.

**Figure 2.**
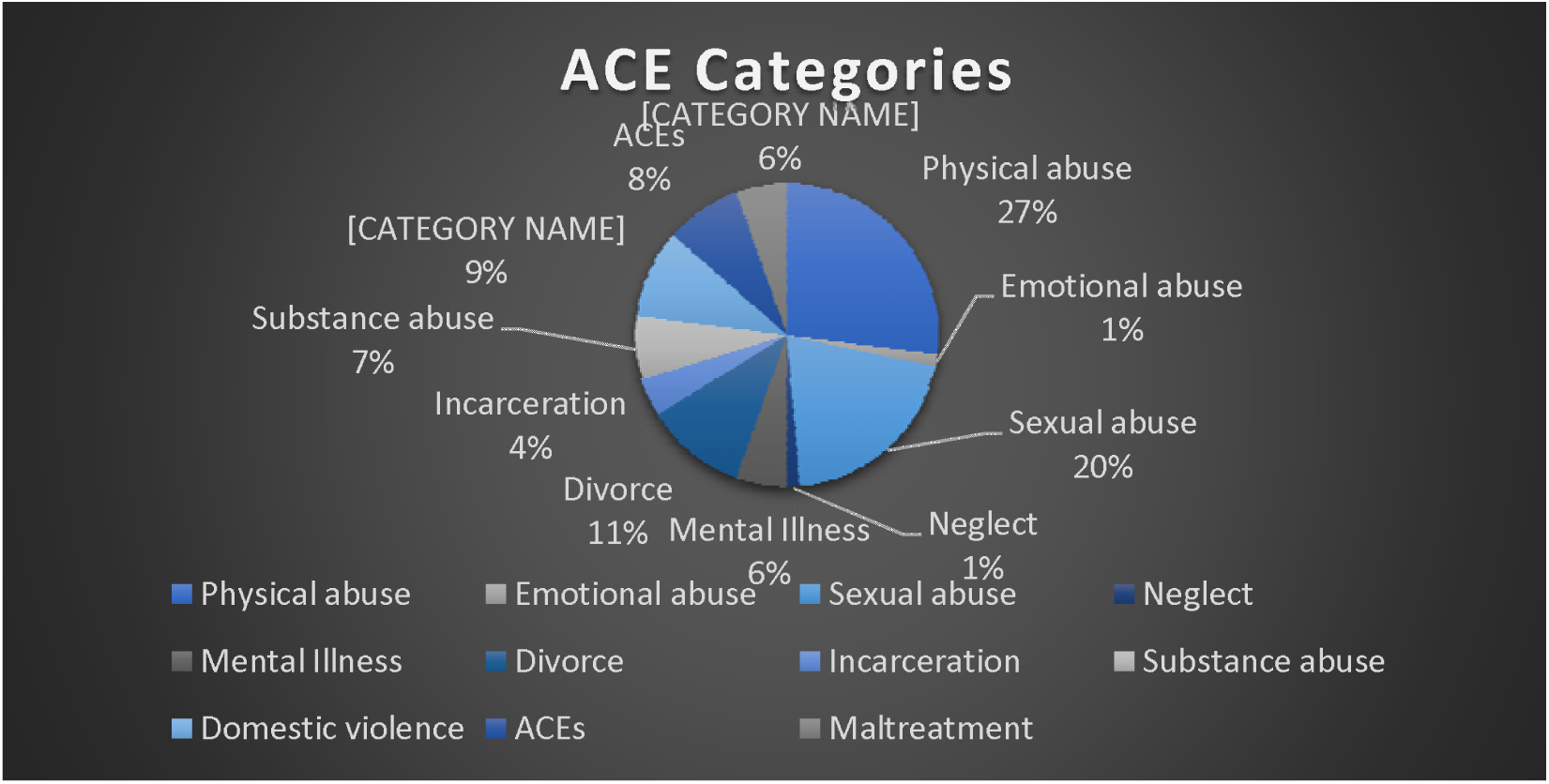
ACE Categories.

Whilst there is a growing body of literature considering siblings experiences of ACEs as a whole, most papers (92%) have taken the approach of exploring individual ACEs rather than the collective. Of the included articles, 107 (72%) directly focussed on siblings whereas the remaining articles explored siblings within their findings despite not being the study’s main focus. Excluding the 37 secondary analysis articles, the most common sample population was children (36%; 53/148) followed by adults providing retrospective reports (17%; 25/148).

The analysis of articles identified three overarching themes: (1) the influence of sibling dynamics, (2) the identification of siblings, and (3) siblings who cause harm.

#### (1) The influence of sibling dynamics

##### The influence of birth order

Exploring the role of the older sibling was a central theme within many articles. Several papers found evidence that older siblings often position themselves as carers, protecting their younger siblings from the ACE, who tend to be seen as more vulnerable (Akerlund, 2017; Callaghan *et al*, 2016; Foroughe & Muller, 2014; Kaye-Tzadok & Davidson-Arad, 2016; Piotrowski, 2011; Ronel & Haimoff-Ayali, 2010; Tedgard, Rastam & Wirtberg, 2019; Vasquez & Stensland, 2015; Woodward & Copp, 2016). Depending on the type of adversity being experienced, the older siblings has been found to either buffer and reduce the potential impact for their younger siblings, or protect them from experiencing further abuse (Foroughe & Muller, 2014; Piotrowski, 2011; Vasquez & Stensland, 2015; Woodward & Copp, 2016). Older siblings have been found to further take on the role of the parent, protecting their siblings and sometimes their parent too. Papers such as Tedgard, Rastam & Wirtberg (2019) refer to this as *parentification*. Children and young people shared with Ronel & Haimoff-Ayali (2010) that due to parental substance abuse and therefore the absence of anyone else taking care of them, older siblings saw themselves as substitutes for their parents.

While a small proportion of articles including Kaye-Tzadok & Davidson-Arad (2016) identified that caring for younger siblings enabled older siblings to find meaning in their abuse, others highlight negative implications such as increased exposure for older siblings, older siblings masking their own needs and older siblings experiencing more negative impacts from the ACE (Al-Quaiz & Raheel, 2009; Callaghan *et al*, 2016; Heins *et al*, 2011; Tailor, Stewart-Tufescu & Piotrowski, 2015). Studies indicate that while older siblings can protect younger siblings from the impact of ACEs, this can be to their own detriment with older siblings holding the burden of responsibility (Buckley, Holt & Whelan, 2007; Carmel, 2019; Tedgard, Rastam & Wirtberg, 2019).

##### The influence of sibling relationships

Several papers have highlighted the influence sibling relationships can have when experiencing ACEs, emphasising their importance (Akerlund, 2017; Frank, 2008; Geschiere, Spijkerma & de Glopper, 2017; Jacobs & Sillars, 2012; Noller *et al*, 2008). This relationship, which for some may be the only viable ongoing relationship in the aftermath of ACEs, can either improve or worsen the impact of their experiences (Kothari *et al*, 2017; Piotrowski, 2011). Following surveys completed with children in Sweden, Jernbro *et al* (2017) identified that children experiencing physical abuse often chose to make their first disclosure to a sibling. Siblings have been found to provide an important source of support, with the absence of a sibling being associated with higher likelihood of experiencing negative effects from ACEs (Geschiere, Spijkerma & Glopper, 2017). Sibling companionship can promote resilience, alleviate potential strains, and predict improved adjustment to the childhood adversity experienced (Jacobs & Sillars, 2012; Vermeulen & Greeff, 2015; Wolfe, 2016).

However, while positive sibling relationships have been found to lessen the impact of ACEs, poor sibling relationships can worsen the impact (Woodward & Copp, 2016). Experiencing childhood adversity can also be the cause of conflict between siblings (Noller *et al*, 2008; Tucker *et al*, 2014; Wolfe, 2016). A case study of two brothers by McGarvey & Haen (2005), for example, describes the siblings as having a ‘traumatic bond’ whereby they would re-enact their abusive experiences with each other. Further, when exploring the impact of divorce, Civitci, Civitci & Fiyakali (2009) recognised that children and young people with a higher number of siblings can feel the need to compete with each other for their parents’ attention, causing conflict within their sibling relationships. Increased conflict and poor sibling relationships have been found to increase levels of loneliness and add further stress in the aftermath of ACEs, worsening the impact (Noller *et al*, 2008; Wolfe, 2016).

A small number of articles have begun to consider sibling relationships with regards to whether they should be placed together having been removed from the care of their parents (Cashmore & Parkinson, 2008; Hindle, 2007; Kothari *et al*, 2017). Hindle (2007) present case studies of siblings within the UK, sharing the decisions made regarding whether to be placed together with their siblings after suffering profound neglect and witnessing violence in the home. While most of these siblings had been place separately, others advocate for siblings to be placed together, arguing it to be critical for the children’s sense of connection, emotional support, and continuity (Cashmore & Parkinson, 2008; Kothari *et al*, 2017).

Davies (2015) is one of the few to have explicitly considered sibling relationships that are not biological, recommending that professionals considering care arrangements should not only consider biological relationships. While most of the articles included in this review either do not specify the type of sibling relationship, or imply a focus on full biological sibling relationships, a small number of articles have given consideration to others like half-siblings and stepsiblings (Gatins, Kinlaw & Dunlap, 2014; Hollingsworth, Glass & Heisler, 2008). Hollingsworth, Glass & Heisler (2008) report that stepsiblings are particularly vulnerable to emotional abuse from parents who may draw siblings in to also cause harm. Gatins, Kinlaw & Dunlap (2014) however found that children and young people affected by divorce appear to be better adjusted when they have half-siblings compared to those who only have full biological siblings. The number of articles explicitly exploring the different types of sibling relationships appear to be lacking when exploring ACEs.

#### (2) The identification of siblings

##### Identifying siblings experiencing ACEs

Some articles, particularly those focussing on maltreatment or neglect, report that when one child has been identified as experiencing abuse there is an increased risk for their siblings to also experience abuse (Hamilton-Gaichritsis & Browne, 2005; Hines, Kantor & Holt, 2006; Lang, Cox & Flores, 2013; MacMillan *et al*, 2013; Witte, Fegert & Walper, 2018). When the childhood adversity results in fatality, surviving siblings are perceived to be at an even greater risk of harm (Damashek & Bonner, 2010). In a retrospective study completed by Witte, Fegert & Walper (2018), around 59% of the 870 sibling pairs reported that both siblings had experienced maltreatment. As well as an increased risk of experiencing the abuse themselves, siblings have also been found to a be at risk of experiencing vicarious trauma from seeing the abuse of their sibling (Hollingsworth, Glass & Heisler, 2008; Keane, Guest & Padbury, 2013). Having a sibling in itself has also been found to be a risk indictor for ACEs, with studies finding those with multiple siblings more likely to experience adversity in their childhood (Benjet *et al*, 2009; Bussemakers, Kraaykam & Tolsma, 2019). Being a twin, furthermore, has been highlighted as increasing risk, with Lindberg et al (2012) believing the increased stress of caring simultaneously for two children is a likely cause of childhood abuse.

Although there is an increased likelihood of multiple siblings experiencing childhood adversity, several papers assert that the traumatic events are often experienced very differently (Boynton, Arkes & Hoyle, 2011; Horn, Hunter & Graham-Bermann, 2013; Skopp *et al*, 2005; Piotrowski, 2011; Piotrowski, Tailor & Cormier, 2014). Even with shared traumatic experiences, children and young people can perceive it very differently and their reactions are unique (Horn, Hunter & Grahamp-Bermann, 2013; Skopp *et al*, 2005). Some articles suggest that age or gender may be the cause of these different perceptions (Piotrowski, 2011; Piotrowski, Tailor & Cormier, 2014). Morrill *et al* (2019) provide other possible causes, including varying exposure, individual perceptions, and unique characteristics such as emotion regulation. With a focus on parental substance abuse, Boynton, Arkes & Hoyle (2011) found that siblings can have different memories of parental alcoholism resulting in one saying their parent was an alcoholic, when the other may not.

##### Implications for service provision

Given the likelihood that when one child experiences abuse siblings will too, a small number of articles have considered what this means for intervention programmes (Ahrons, 2007; Farnfield, 2017; Morrill *et al*, 2019; Renner & Boel-Studt, 2017; Renner & Driessen, 2019; Skopp *et al*, 2005). Some studies have argued that interventions are more meaningful when they include siblings, however in most cases only one child from a family will be referred (Ahrons, 2007; Farnfield, 2017). Recommendations have been made that service providers would be wise to assess all siblings within a family when one has experienced ACEs, and siblings should furthermore be considered when planning services, completing screenings and undertaking assessments, (Morrill *et al*, 2019; Renner & Boel-Studt, 2017). Renner & Driessen (2019) go further to believe that while siblings need increased attention from services, the needs of all family members should be assessed. Skopp *et al* (2005) are clear that in practice this does not simply mean referring all siblings to support programmes, as this would not be a good use of resource. Rather, individually assessing all siblings within families is much more appropriate.

#### (3) Siblings who cause harm

##### Sibling violence and abuse

Substantial consideration has also been given to siblings causing harm to other siblings, with this being the focus of nearly 40% of included articles. Several of these papers have identified abuse from a sibling as one of the most common forms of violence in the home (Finkelhor, Ormrod & Turner, 2009; Kim & Kim, 2019; McDonald & Martinez, 2016; Meyers, 2014; San Kuay *et al*, 2016; Soler *et al*, 2015). Multiple papers however highlight a significant difference in the perception and understanding of this form of abuse; despite its prevalence and documented detrimental impact, sibling violence and abuse is often dismissed as normal or expected (Button & Gealt, 2010; Desir & Karatekin, 2018; Kim & Kim, 2019; Phillips *et al*, 2018; Sporer, 2019; Tippett & Wolke, 2015; Van Berkel, Tucker & Finkelhor, 2018). Parents in particular have been highlighted as holding these views, being found to often minimise the frequency, severity and impact (Tucker *et al*, 2013). Aggression between siblings can often be seen as normative and harmless by parents (Miller *et al*, 2012; Van Berkel, Tucker & Finkelhor, 2018) with some having difficulty in determining what behaviours are acceptable (McDonald & Martinez, 2019). Tompsett, Mohoiney & Lackey (2018) further find that children themselves can also minimise abuse and violence from a sibling.

Hoffman, Kiecolt & Edwards (2005) and Tompsett, Mohoiney & Lackey (2018) are in the minority, having given consideration to theoretical understandings of sibling violence and abuse. They highlight from a social learning perspective that children who experience abuse from a parent are likely to then use violence against a sibling (Hoffman, Kiecolt & Edwards, 2005). Aggressive parents model this behaviour as acceptable, leading to their children becoming more aggressive themselves (Thompsett, Mohoiney & Lackey, 2018).

##### Sibling sexual abuse

Within the dataset of articles exploring sibling violence and abuse are a subset of papers which have explored children and young people who are sexually abused by a sibling; a crossover between the sibling violence and abuse articles and the sexual abuse articles (see figure 3). Many articles share the detrimental impact this ACE can have, finding survivors to experience anxiety, depression, and posttraumatic stress disorder, as well as being estranged from siblings or parents, and having potential future relationship or intimacy problems (Beard *et al*, 2013; Carlson, 2011; Falcao *et al*, 2014). Of significance are some of the unique implications of having a sibling cause this harm. Studies have found that it is possible for the abuse to be longer-lasting due to the accessibility of siblings, and even small age gaps between siblings can create a significant power imbalance (Carlson, Maciol & Schneider, 2006; Tener & Silberstein, 2019). Tener & Silberstein (2019) further highlight that for many, this abuse only ends when the siblings leaves the family home.

**Figure 3.**
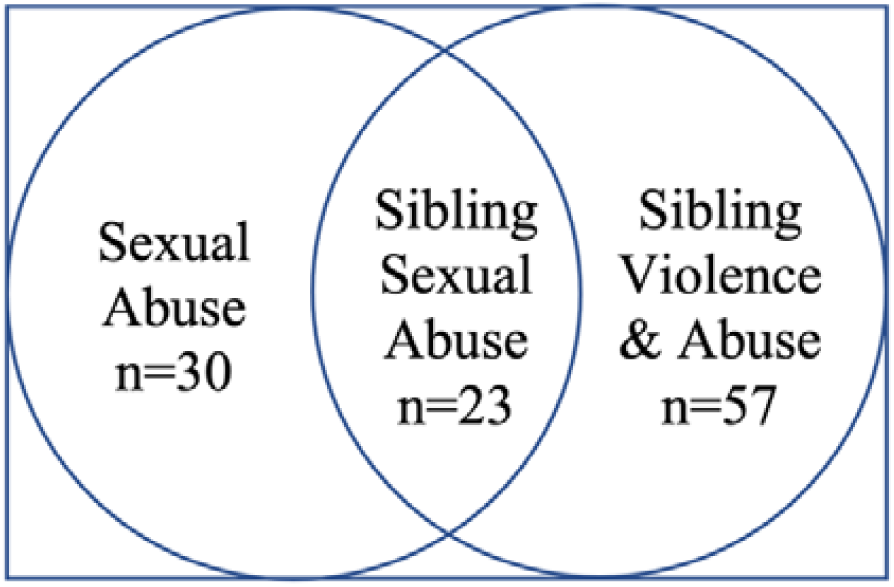
Venn Diagram.

Like with other forms of sibling abuse, studies suggest that sibling sexual abuse is often normalised, most often by parents and guardians (Tarshish & Tener, 2019; Tener *et* al, 2018). Some parents have been found to see these behaviours as harmless, perceiving it as a form of age-appropriate curiosity; others are believed to minimise it or deny due to shame (Tarshish & Tener, 2019; Tener *et* al, 2018; Tener *et al*, 2019). Bass *et al* (2006), for example, present two contrasting case studies of families where there has been sibling incest; one of the families found the behaviour as normal whereas the other did not. Although there is a high prevalence of sibling sexual abuse, this ACE has been found to be rarely reported (Carlson, Maciol & Schneider, 2006; Celbis, Ozcan & Ozdemir, 2006; Joyal, Carpentier & Martin, 2016; Katz & Hamama, 2017). Some articles have argued this to be due parental perceptions normalising the behaviour, or not recognising the serious nature of the abuse, while others have found parents to respond to disclosures with disbelief (Krienert & Walsh, 2011; McDonald & Martinez, 2017; Morrill, 2014).

This underreporting has been argued to have hindered large-scale research in this area (Krienert & Walsh, 2011). One of the difficulties associated with researching sibling sexual abuse is the lack of a universally accepted definition and understanding (Caffaro & Conn-Caffaro, 2005). Beard *et al* (2013), for example, define this abuse as any form of sexual behaviour among relatives in their own study. Bass *et al* (2006 p.93) in comparison defines it as “sexual behavior between siblings that results in feelings of anger, sadness, or fear in the child who did not initiate the behavior”; implying that some sexual behaviour would not be considered abusive. These different understandings means that those included in the sample population can vary significantly, potentially affecting the findings of studies.

## Discussion

This scoping review has provided detailed insight into what is currently known about the experiences of siblings when living with ACEs. The PAGER framework (Bradbury-Jones *et al*, 2021) has been utilised within this review as a means of offering a structured approach to its analysis and reporting. Five main patterns have been identified within this review which will now structure the discussion, where advances and gaps will be presented along with evidence for practice and research recommendations. A full overview of this framework is provided in Table 1.

**Table 1.** PAGER Framework.

| **Table 1 – PAGER Framework** |  |  |  |  |
| --- | --- | --- | --- | --- |
| **Patterns** | **Advances** | **Gaps** | **Evidence for Practice** | **Research Recommendations** |
| The influence of birth order | There is evidence of older siblings protecting younger siblings from the impact of ACEs | There is the need for ongoing empirical work exploring the potential increased impact on older siblings who are shielding younger siblings from the ACE | It is important that support services consider their response to older siblings who may take far more burden than their younger siblings | To expand research exploring the potential increased impact on older siblings who are shielding younger siblings from the ACE |
| The influence of sibling relationships | Understanding is growing around how a positive sibling relationship can reduce the impact of ACEs | Little consideration has been given to the different types of siblings (full, step, half), and whether this influences the impact of ACEs | It is important for statutory bodies to consider the positive benefit of sibling bonds, especially when making decisions around whether to place siblings together when removed from the care of their parents. | Far greater attention needs to be given to research exploring different sibling relationships (full, half, step) |
| Identifying siblings experiencing ACEs | There is some evidence that when one sibling experiences ACEs, it is likely that other siblings are also experiencing it | There is limited evidence about how agencies identify whether other siblings have also experienced the ACE | It is important to consider all siblings when one has presented with experiencing childhood adversity. Siblings need to be included in screening and referrals into interventions | Research is needed that explores how agencies identify whether other siblings have also experienced ACEs |
| Siblings who cause the harm | Substantial consideration has been given to siblings harming other siblings.<br>The abusive behaviour is often normalised and minimised when it is a sibling causing the harm | Focus tends to be on parents minimising the abuse, with little consideration being given to the views and responses of practitioners and services | It is important that support services consider their response to sibling abuse, especially when the physical abuse can be bi-directional. | Need more research which considers the views and responses of practitioners and services to children and young people causing harm to siblings |
| Focus on individual ACEs | There is a growing body of literature considering siblings experiencing ACEs.<br>Most explore individual ACEs rather than the collective. | Some ACEs are explored substantially more than others.<br>More than half do not offer any theoretical understanding. | More evidence will emerge from future research considering theory | Studies are required that explore the ACEs currently given less attention.<br>Theoretical understanding is vital within this area of research |

### (1) The influence of birth order

A clear advance identified within this review has been the overwhelming evidence of older siblings being a protective factor for their younger siblings, shielding them from adverse experiences. Described by Tedgard, Rastam & Wirtberg (2019) as *parentification*, their buffering role also finds them taking on the role of the parent. Whilst there has been lot of recognition for the vulnerabilities of younger siblings, substantially less thought has been given to older siblings. Further consideration is now needed around how this protective role impacts the older sibling. This knowledge will be of significance to frontline support services who need to consider their response to older siblings who may take far more burden than their younger siblings.

### (1) The influence of sibling relationships

One of the gaps evident from this scoping review is the lack of research exploring sibling types outside of biological siblings. Whilst there is a wealth of evidence around both the positive and negative influence sibling relationships can have, this has primarily been done from the sole perspective of biological siblings. Very few studies have explored step-, half- and adoptive-siblings; the majority either do not specify the type of sibling relationship, or imply a focus on full biological sibling relationships. The findings of Gatins, Kinlaw & Dunlap (2014) that divorce appear affect those with half-siblings differently to those who only have full biological siblings, indicate that this is an area that also needs more consideration. Further research needs to be undertaken which explores the wide range of sibling relationships within society in order to understand if this influences the impact of ACEs. It will be important for statutory bodies to consider these findings in practice, especially when making decisions around whether to place siblings together when removed from the care of their parents.

### (2) Identifying siblings experiencing ACEs

Patterns and gaps have also been identified within this review relating to the identification of siblings experiencing ACEs. There is growing evidence that when one sibling experiences adversity, it is likely that their other siblings are also experiencing it, or be at risk of experiencing vicarious trauma. Lindberg *et al* (2012) identified twins in particular as being at increased risk of experiencing childhood abuse. Studies recommend the assessment of all siblings within a family when one has experienced childhood adversity, while remaining clear that this does not simply mean referring all siblings to support programmes, as this would not be a good use of resource (Morrill *et al*, 2019; Renner & Boel-Studt, 2017; Skopp *et al*, 2005). Knowledge of how agencies approach the identification of siblings, however, is lacking meaning further targeted research on around this is required.

### (3) Siblings who cause the harm

Substantial advances have been made in the exploration of siblings harming other siblings. An overwhelming pattern across the studies is how this abusive behaviour is often normalised and minimised, especially by parents. Considerable underreporting has been highlighted as hindering research in this area (Krienert & Walsh, 2011), with little consideration being given to the views and responses of practitioners and frontline services. Tippett & Wolke (2015) recognise the unique nature of this childhood adversity in that the abusive behaviour can be bi-directional, meaning the response of support services is of importance. With the current research focussing more on the views of parents, further research is needed which considers the views and responses of practitioners and services to children and young people causing harm to siblings.

### (4) Focus on individual ACEs

Whilst some advances are being made in terms of research exploring ACEs as a collective, the majority have explored them in isolation. This has resulted in some types of childhood adversity, such as physical abuse and sexual abuse, being explored to a greater degree. Research exploring some of the less studied ACEs, or ACEs as a collective, is required to develop current understanding. Children and young people who experience one form of childhood adversity are likely to experience multiple, meaning exploring ACEs as a collective would be appropriate.

## Limitations

Limitations of this review include the need to reduce the timeframe in which we included articles. Whilst we had planned to include articles from 1995-2019 to capture all articles published since the first ACEs study was conducted, this timescale was reduced by 10 years due to not having the resource available to review the high volume of articles, meaning the final agreed timescale was 2005-2019 and a further 84 articles were excluded at this stage. The current study was also limited to empirical articles published in peer-reviewed journals, but we recognise that there may be additional knowledge outside of this that was not available through our search mechanisms.

## Conclusion

This scoping review has examined what is currently known about the experiences of siblings living with ACEs. Included articles have highlighted overwhelming evidence of older siblings shielding younger siblings, and the likelihood that when one sibling experiences adversity, other sibling will be experiencing it themselves or vicariously. The implications of this in practice are that support services and statutory bodies need to ensure considerations are being made for all siblings when one has presented with experiencing childhood aversity, especially older siblings who make take far more burden. Given that more than half of the included articles did not offer any theoretical understanding, this area is of significant importance for future research. Far greater attention is also needed for research exploring different types of sibling relationships (full, step, half), and whether they influence the impact of ACEs have on children and young people.

## Data Availability

This is a scoping review and a list of all included articles is available upon reasonable request to the authors

